# Spontaneous Cervical Artery Dissection: A Case Series

**DOI:** 10.1101/2020.08.25.20181933

**Authors:** Amedra Basgaran, Catherine Hsiao-wei Hsu, Aravinth Sivagnanaratnam

## Abstract

Cervical artery dissection refers to dissection of the vertebral or carotid arteries, and accounts for up to 20% of strokes in those under 45. Antithrombotic therapy is therefore essential to limit thrombosis at the site of injury and distal neuronal damage. However, the exact choice of drugs, timing and duration of therapy remain a challenging decision.

A review of data was conducted on three cases of unprovoked cervical dissection at our stroke center between 2017 and 2020. They include bilateral internal carotid artery dissection, right vertebral artery dissection, and left internal carotid artery dissection.

Three key outcomes were identified: narrowing, occlusion and pseudoaneurysm; such outcomes dictated our management approach. Two patients were given antiplatelet therapy for at least one year. The patient with bilateral dissection was perceived to have higher thromboembolic risk, due to the presence of a free-floating thrombus. Thus, he was anticoagulated for a year, and thereafter given antiplatelet therapy.

The evidence behind long-term management of carotid artery dissection remains equivocal. There is no strong evidence favouring anticoagulation over antiplatelets or vice versa. Anticoagulation tends to be preferred in cases of severe stenosis, occlusion or pseudoaneurysm, to reduce the risk of thromboembolic phenomena. Anti-platelets are preferred where there is a high risk of haemorrhagic transformation or contraindication to anticoagulation. The duration of secondary prevention is similarly unclear. Advances in radiology and increased follow-up have resulted in uncertainty on the management of incomplete healing at the six-month point. Varying clinical practice has been identified and there is a lack of a clear guideline. We propose continuing antithrombotic therapy in cases of incomplete healing, as in our case series. Nevertheless, we require more data on the subject and thus suggest an initial nation-wide survey to compare the different management strategies followed by large-scale retrospective analyses comparing long-term outcomes.

## Introduction

Cervical artery dissection (CAD) refers to dissection of the vertebral or carotid arteries. It accounts for up to 20% of strokes in those under 45[1] due to association with a high risk of intracranial microembolisms[2]. It thus has potentially devastating consequences, including neuropathy, ischaemic stroke, subarachnoid haemorrhage and death. Antithrombotic therapy is therefore essential to limit thrombosis at the site of injury and distal CAD-related neuronal damage. However, the exact choice of drugs, timing and duration of therapy remain a challenging decision. Furthermore, identification of underlying risk factors can assist with reducing further risk of dissection; yet there lacks a clear guideline for such investigations.

## Methods

A review of data was conducted on three cases of cervical dissection at our stroke centre between 2017 and 2020. We compare and contrast their presenting and imaging features, management approaches and outcome. All three patients have been followed up at 6 monthly intervals, with computed tomography angiography performed to assess progress. Written informed consent was obtained from all patients for their anonymized information to be published in this case series.

## Results

All cases were unprovoked and involved either the carotid or vertebral artery. Presenting features varied based on vascular territory affected, and are described in Table 1. The cases include bilateral internal carotid artery dissection, right vertebral artery dissection, and left internal carotid artery dissection.

**Table 1.** Presenting features, CT findings, outcome and management plans of three patients presenting with cervical artery dissection at our centre.

| **Patient:** | **1** | **2** | **3** |
| --- | --- | --- | --- |
| **Presentation** | <u>1st dissection:</u> Sudden onset headache and neck pain.<br>Temporary right sided arm weakness<br><u>2nd dissection:</u> Sudden onset headache and neck pain. Temporary left sided arm weakness. | Sudden onset vertigo and vomiting, resolved after 3 days. | Sudden onset left blurred vision, right hand numbness, expressive dysphasia, unsteadiness. |
| **CT Angiogram Findings** | Left internal carotid artery dissection<br>Right internal carotid artery dissection | Right vertebral artery dissection<br>Right cerebellar infarct | Left internal carotid artery dissection<br>Left middle cerebral artery territory infarct |
| **Co-Morbidities** | Borderline hypertension | Meningitis 5 years prior to event.<br>Smoker | Smoker |
| **Outcome at 6 Months** | Left pseudoaneurysm<br>Right narrowing | Occlusion | Narrowing |
| **Outcome on Latest Review** | 2 years: Left pseudoaneurysm, Right recanalisation | 1 year: Occlusion | 6 months: Narrowing |
| **Management** | Warfarin for 1 year<br>Clopidogrel 75mg ongoing | Aspirin 300mg for 2 weeks | Aspirin 300mg for 2 weeks |
|  |  | Clopidogrel 75mg<br>ongoing | Clopidogrel 75mg<br>ongoing |

**Table 2.** Risk Factors identified for Carotid Artery Dissection and Investigations performed to rule out such causes.

| Category | Risk Factors | Salient Negative Investigations |
| --- | --- | --- |
| **Autoimmune** | Antiphospholipid Syndrome<br>Vasculitides | Antiphospholipid antibodies<br>ANCA, ANA, ENA, RhF, ESR |
| **Collagenous** | Ehlers-Danlos Syndrome<br>Marfan's disease<br>Collagen vascular disease | Genetic testing |
| **Structural** | Autosomal polycystic kidney disease<br>Fibromuscular dysplasia | Renal Ultrasound<br>CT angiogram |
| **Toxin** | Cocaine-induced vasospasm | Urine toxicology screen |

The investigation of underlying risk factors was taken further with patient 1, given his bilateral nature. This patient had two risk factors identified: borderline hypertension and a habit of “cracking his neck”. Although there are no reports of this mechanism, there are reported cases of carotid artery dissection following cervical manipulation coupled with an underlying arteriopathy such as idiopathic cystic medial degeneration[3], and minor trauma such as coughing [4]. This habit coupled with his borderline hypertension is a potential but weak hypothesis explaining his dissection.

There are 3 key outcomes identified in cervical artery dissection: narrowing, occlusion and pseudoaneurysm; all of which are demonstrated in this case series. Such outcomes have dictated our management approach. Two patients were given antiplatelet therapy for at least one year. The patient with bilateral dissection was perceived to have higher thromboembolic risk, due to his bilateral nature and also the presence of a free-floating thrombus. Thus, he was anticoagulated with warfarin for a year, and thereafter given antiplatelet therapy.

## Discussion

Given the rarity of bilateral spontaneous carotid artery dissection, we proceeded to investigate for other underlying conditions that could have predisposed this patient to such a pathology (Table 1). We have comprised a list of risk factors investigated for and grouped them according to category: autoimmune causes such as vasculitis of any origin, collagenous causes such as Marfan’s syndrome, structural causes such as autosomal polycystic kidney disease, and toxin-mediated causes such as cocaine induced vasospasm. All investigations have come back as negative.

Genetic testing was negative for collagen vascular disease. A variant of unknown significance was identified in the FBN2 gene on the FTAAD (familial thoracic aorta and dissection) gene panel. This was thought unlikely to be related to the carotid artery dissection as such mutations are usually associated with Beals syndrome (congenital contractural arachnodactyly).

A literature search identified further conditions to consider, namely osteogenesis imperfecta and hyperhomocysteinemia[5].Furthermore, there are reported cases where predisposing risk factors are yet to be defined as conditions, such as carotid artery tortuosity identified on CT angiogram imaging[6].

Cervical artery dissection is treated with anti-platelets or anti-coagulation, with no evidence favouring one[7,8,9]. A number of meta-analyses [10,11] as well as a powerful multicenter prospective RCT (CADISS) [12] found no significant difference between the two therapies. The literature suggests anticoagulation in cases of severe stenosis, occlusion or pseudoaneurysm, on the basis that they reduce the risk of thromboembolic phenomena[7,8]. In contrast, antiplatelets are preferred in patients with a poor prognosis or large infarcts, to reduce the risk of haemorrhagic transformation[7,9,13].

The suggested time-length for treatment is more unclear. Radiographic improvement in cervical artery dissection largely takes place in the first 6 months[14], suggesting a similar time length for treatment[7]. Yet, this timeline is also confounded by the varying complications that can occur, such as narrowing of the artery or the formation of a pseudo-aneurysm. In such cases where there is incomplete healing at the six-month point, there are no trials or clear guideline on how long to treat with anti-thrombotic therapy. A small survey of clinicians at the national UK Stroke Forum in 2019 on their management of cervical artery dissection similarly reflected varying practice (Table 3). The majority preferred antiplatelet therapy, with some clinicians reportedly using dual antiplatelet therapy. Anticoagulation was preferred in cases where there was a free-floating thrombus. The duration of therapy also varied with some clinicians opting for 6 months, and some opting for lifelong therapy especially when using antiplatelets. Such variation in practice highlights gaps in knowledge requiring rectification.

**Table 3.** Survey of Management of Cervical Artery Dissection by Clinicians at UK Stroke Forum 2019

|  | **<u>Preference on Therapy</u>** | **<u>Factors affecting Choice of Therapy</u>** | **<u>Duration of therapy</u>** |
| --- | --- | --- | --- |
| **1** | Anticoagulant | Anticoagulation if ischaemic changes on imaging and young patients. | 6 months |
| **2** | Antiplatelet | Anticoagulation if free-floating thrombus present. | Lifelong if antiplatelet, 6 months if anticoagulant |
| **3** | Antiplatelet | Antiplatelet if young and no comorbidities. Anticoagulation if older with ischaemic heart disease. | 6 months |
| **4** | Antiplatelet | Unsure | Unsure |
| **5** | Antiplatelet | Unsure | Unsure |
| **6** | Antiplatelet | Unsure | Lifelong |
| **7** | Antiplatelet | Initial antiplatelet therapy. If no healing at 6 months, anticoagulation. | 6 months |
| **8** | Antiplatelet | Anticoagulation if free-floating thrombus present. | 6 months |
| **9** | Antiplatelet | Anticoagulation if comorbidities resulting in pro-coagulable state. | Lifelong |
| **10** | Both | Unsure | Unsure |

## Conclusion

This case series has provided us with valuable learning points on how to investigate and treat patients with spontaneous carotid artery dissection. The evidence behind long-term management of carotid artery dissection however remains equivocal. There is no strong evidence favouring anticoagulation over antiplatelets or vice versa. Anticoagulation tends to be preferred in cases of severe stenosis, occlusion or pseudoaneurysm, on the basis that anticoagulation reduces the risk of thromboembolic phenomena. Anti-platelets are preferred where there is a high risk of haemorrhagic transformation or contraindication to anticoagulation, and are a more convenient choice. Furthermore, the duration of secondary prevention is unclear. Advances in radiology and increased follow-up have resulted in uncertainty on the management of incomplete healing at the six-month point. Varying clinical practice has been identified and there is a lack of a clear guideline. We propose continuing antithrombotic therapy in cases of incomplete healing, as in our case series. Nevertheless, we require more data on the subject and thus suggest an initial nation-wide survey to compare the different management strategies, particularly looking at duration of therapy, followed by large-scale retrospective analyses comparing the long-term outcomes associated with these different practices.

## Data Availability

Not applicable.

## Acknowledgements

Nil.

---

**Figure 1.**
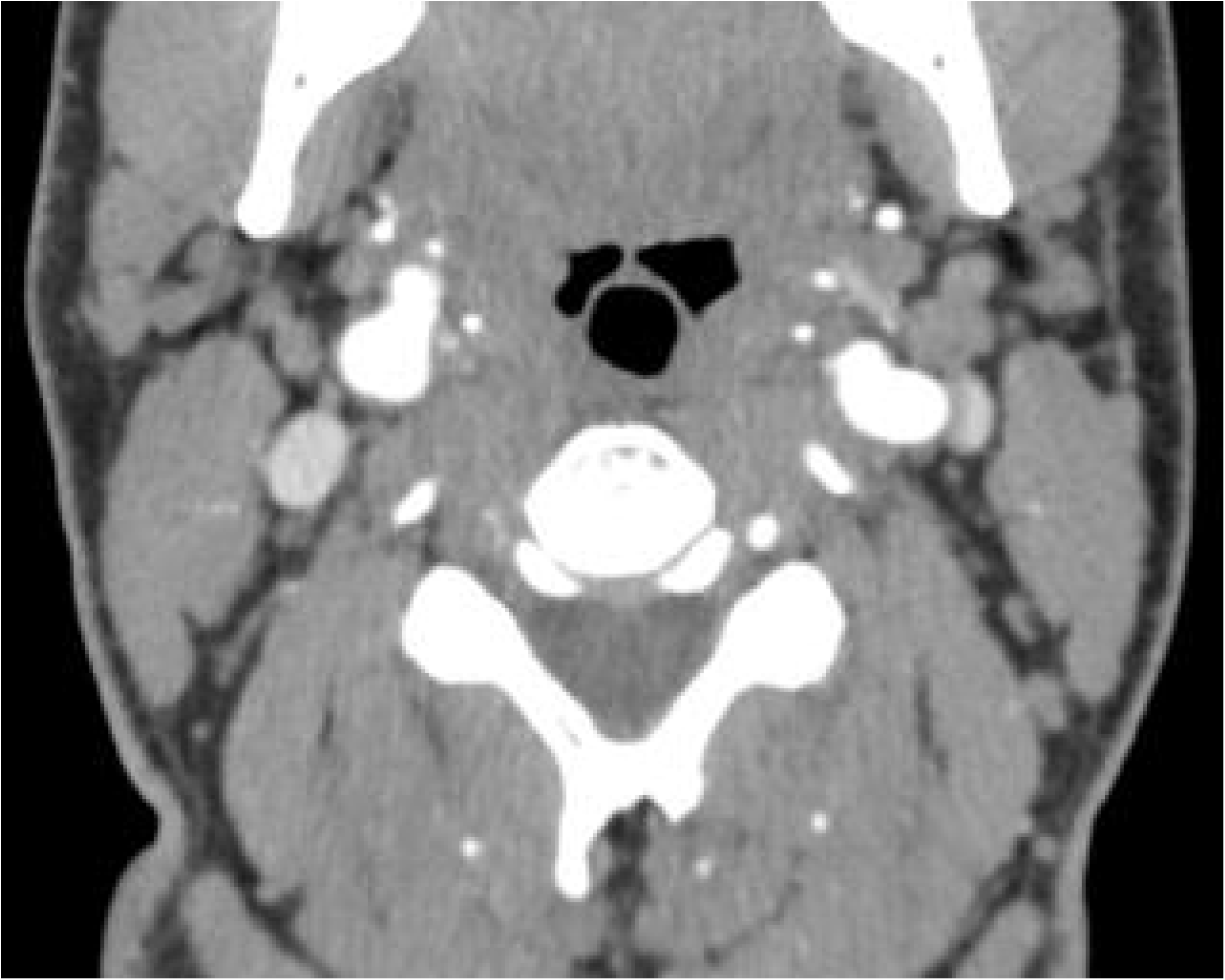
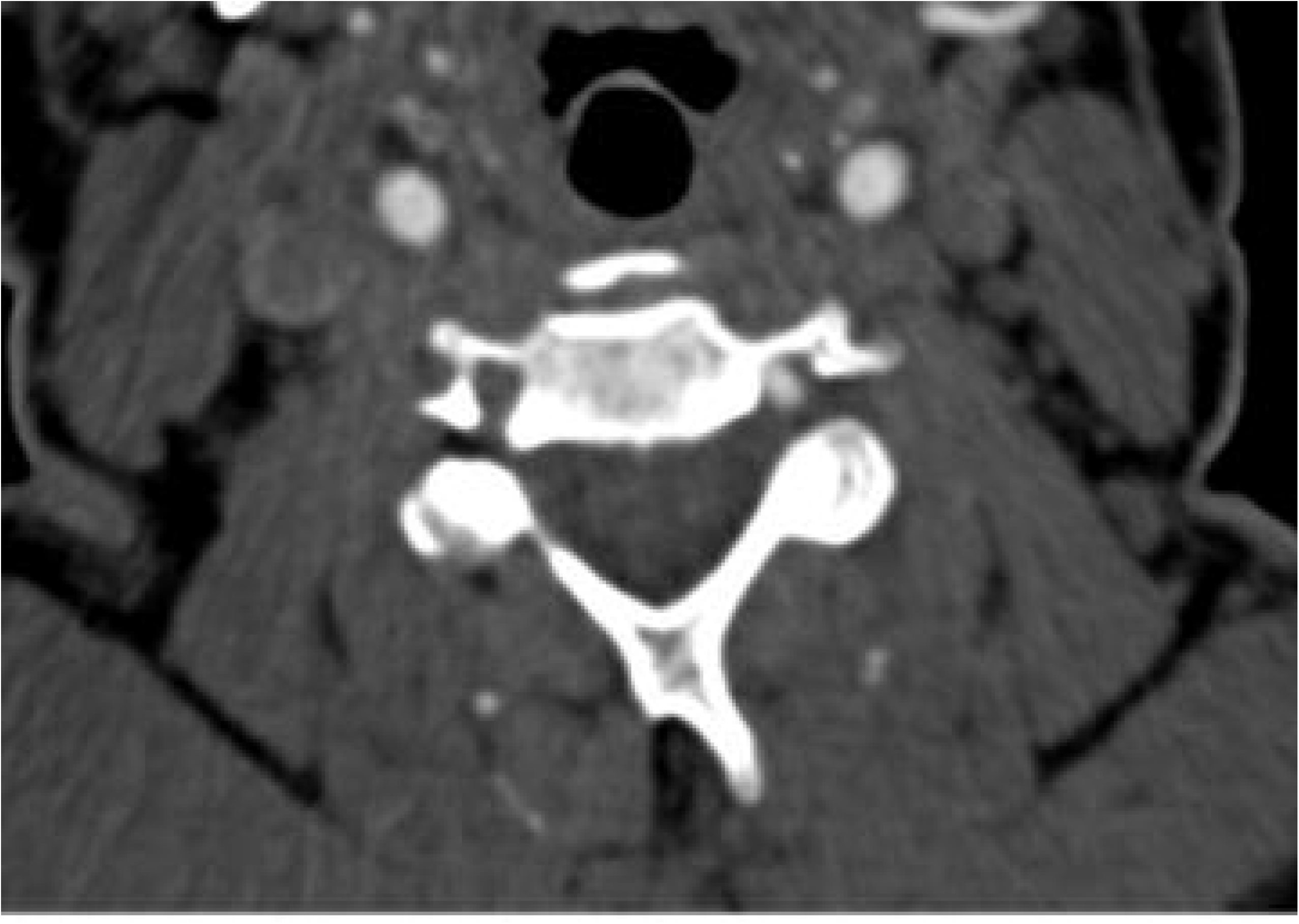
Example CT Imaging of Patient 2 **A**, Initial CT Head identifying Right Vertebral Artery Dissection **B**, Follow Up CT Head at 6 Months identifying Right Vertebral Artery Occlusion

**Figure 2.**
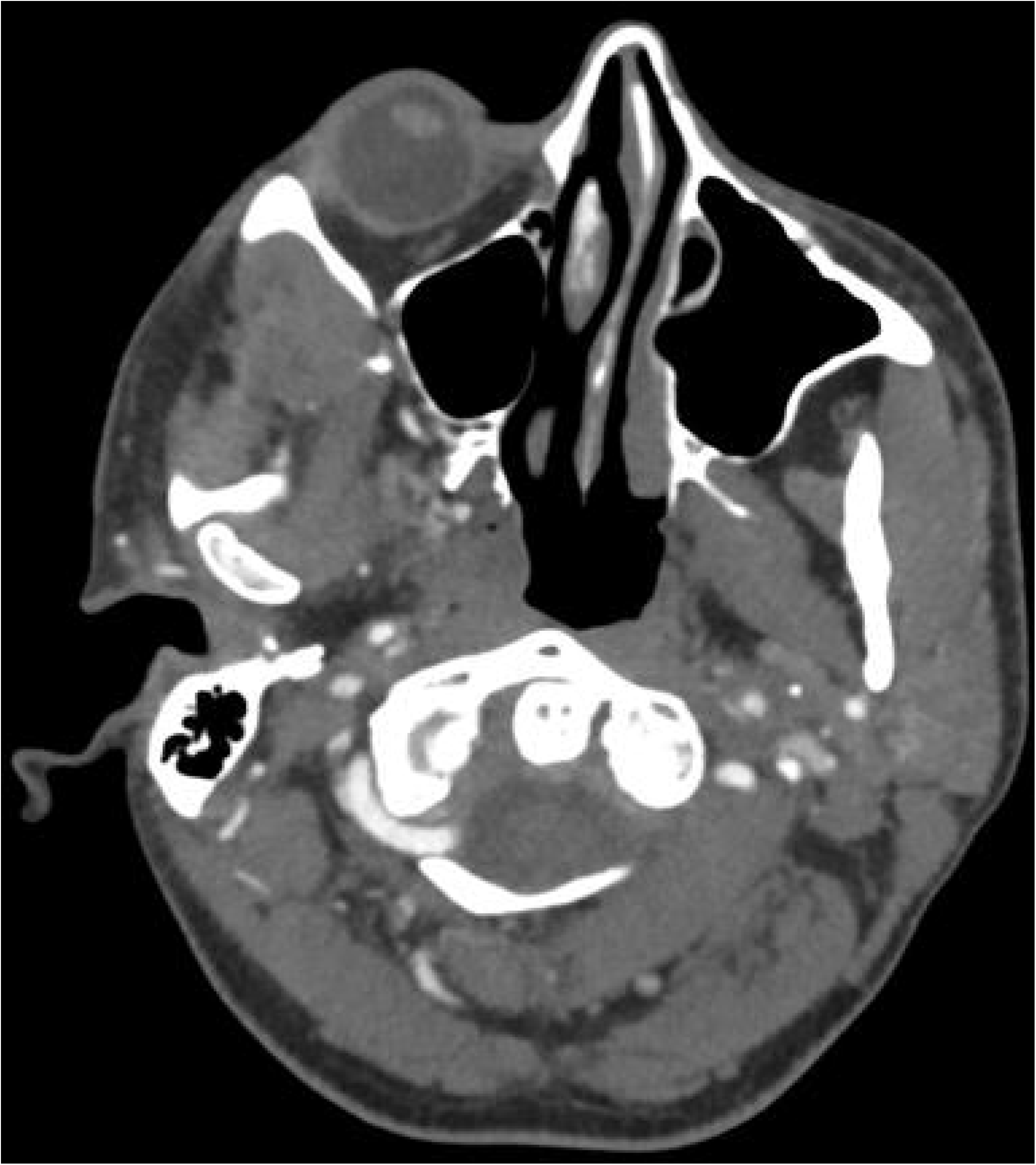
CT Head of Patient 1 in Axial Plane identifying Left Carotid Artery Dissection Flap and Right Carotid Artery Narrowed Lumen with a Mural Haematoma

**Figure 3.**
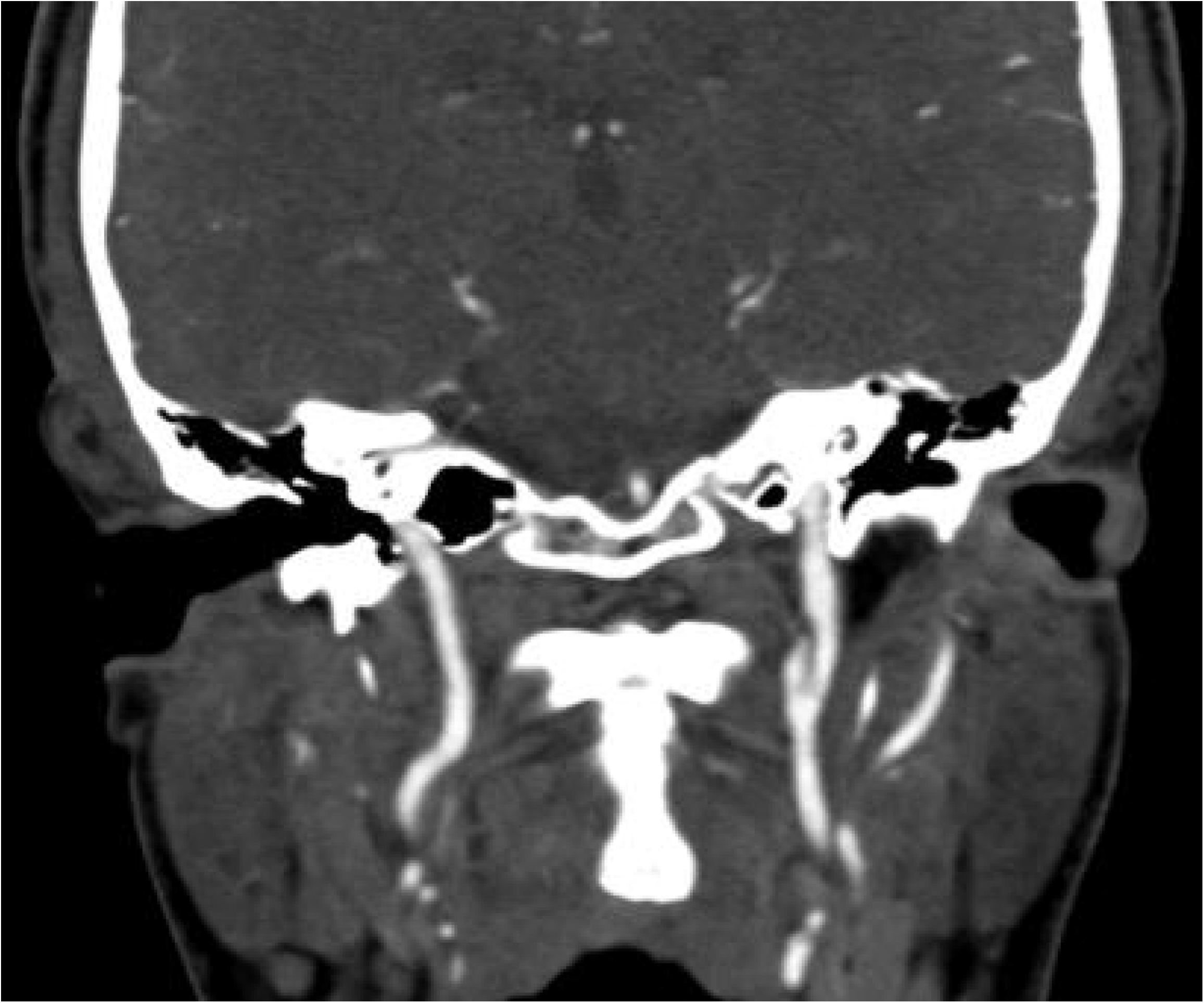
2 Year Follow up CT Head of Patient 1 in Coronal Plane identifying Left Carotid Artery Pseudoaneurysm and Right Carotid Artery Recanalisation

## Online Supplement

Nil

